# Biologically informed geometry and force distribution improve task performance in agonist/antagonist tendon-driven prosthetic hands

**DOI:** 10.64898/2026.04.06.26350199

**Authors:** Lorena Velásquez, Jeremy D. Brown

## Abstract

Prosthetic devices balance functionality and usability to support activities of daily living (ADLs). However, many designs rely on rigid end effectors that, while anthropomorphic in form, lack biomimetic design principles. This mismatch increases cognitive and physical burden, reducing adoption rates. We developed the Human-inspired Actuator Modeling and Reconstruction (HAMR) process, a user-centered framework informed by individual morphology and functional needs, to generate customized agonist/antagonist tendon-actuated end effectors. Using HAMR, we created the Tendon Actuated Prosthetic Hand (TAPH), which integrates human-derived geometry with adaptive force distribution to promote natural object interaction. In a study with 12 participants without limb difference, TAPH was compared to a structurally similar tendon-actuated hand with generalized anthropomorphic geometry across three ADL tasks of varying complexity. TAPH significantly improved task performance and reduced physical effort, mental workload, and frustration, particularly during gross motor tasks. For fine motor tasks, performance improved under stable conditions but not during tasks requiring dynamic precision and continuous coordination. These findings highlight the functional benefits of biologically informed prosthesis design and support biomimetic principles in enhancing performance and user experience.

## Introduction

Upper-limb prosthetic hands are widely used to support activities of daily living (ADLs), yet their functional performance varies substantially across tasks and users. Existing designs range from simple body-powered hooks to multi-articulating myoelectric hands, many of which emphasize generalized grasping capability rather than task-specific interaction^1,2^. Despite advances in actuation, sensing, and control, achieving efficient and intuitive manipulation across both gross and fine motor tasks remains a persistent challenge for prosthesis users^3,4^.

Beyond actuation, commercial prosthetic hands are designed to facilitate interaction with the environment through complex grasping capabilities, but often lack the level of customization required to meet the unique needs of individual users^3^. Prosthetic device paradigms have deviated from user-centered design principles, resulting in expensive, generic, mass-produced devices that are often uncomfortable, unreliable, and both physically and cognitively demanding to use^5–7^. To that end, recent research has begun investigating the utility of modular hands that employ task-specific mechanisms for various activities of daily living^8^. Still, these solutions may not always be practical for everyday use by the average user. As a result, many unilateral upper-limb prosthesis users rely on their natural limb for dexterous manipulation and employ the prosthesis primarily in a stabilizing or supporting role rather than as the primary manipulator^4^.

The reliance on the prosthesis as a secondary or supportive manipulative tool arises in part from limitations in traditional prosthetic hand designs that constrain how the device adapts to object interaction. Traditional designs are rigid and stiff, relying on control strategies that prescribe specific joint positions and tightly couple hand aperture with hand impedance, the effective stiffness or compliance of the hand. In these systems, interaction forces are largely determined by predefined control parameters, thereby limiting the hand’s ability to adapt to variability in object shape or contact conditions. Tendon-driven prosthetic hand designs have been proposed as an alternative to these fixed-impedance mechanisms. By routing actuation forces through tendons, these designs can decouple hand impedance from aperture and joint position. This decoupled structure enables the hand to conform to object geometry and adjust contact forces during interaction in a manner more consistent with biological hand function^9–12^. Importantly, tendon-driven prosthetic hands have been shown to outperform traditional commercially available prosthetic hands in functional task performance and user efficiency, particularly in activities requiring adaptive grasping and force modulation^8,13^.

This prior work has demonstrated that underactuated tendon-based architectures can support stable grasps across diverse object morphologies and task demands^14,15^. However, many tendon-driven systems rely on generalized anthropomorphic geometries that are not explicitly aligned with human hand biomechanics^16,17^. Prosthetic hand geometry directly influences grasp formation, contact stability, and compensatory movement strategies. Finger proportions, joint axis placement, thumb orientation, and palmar geometry affect how forces are transmitted during manipulation and how users coordinate motion during interaction^2^. Despite these biomechanical considerations, it remains unclear whether biologically informed hand design improves functional task performance or reduces user workload in tendon-based architectures. The specific contributions of human-derived geometry and adaptive force distribution have not been evaluated under matched control conditions.

To enable controlled evaluation of biologically informed hand geometry, we developed the Human-inspired Actuator Modeling and Reconstruction (HAMR) process, a modeling and fabrication workflow that derives prosthetic hand geometry directly from human anatomy. HAMR integrates physical molding, three-dimensional scanning, and computer-aided design to capture hand morphology, define anatomically aligned joint axes, and establish tendon routing paths that are consistent with the scanned geometry. This process enables the creation of tendon-driven prosthetic end effectors whose finger proportions, joint locations, and thumb placement are explicitly mapped from a human hand, while remaining compatible with conventional tendon actuation and myoelectric control strategies.

Using the HAMR process, we created the Tendon Actuated Prosthetic Hand (TAPH), a tendon-driven end effector that integrates biologically derived hand geometry with an adaptive force distribution architecture informed by human grasping behavior. In this study, we evaluate the functional impact of this combined hand design through a comparative assessment of two agonist/antagonist tendon-driven prosthetic end effectors: TAPH and a structurally similar hand with a generalized anthropomorphic geometry and more traditional tendon coupling, the Bionic Skeletal Hand (BSH)^13,18^. Both devices employ identical motors, control logic, and user interfaces, enabling differences in task performance and user workload to be attributed to hand-level design features, including geometry and force distribution, rather than differences in control strategy. Performance was evaluated in participants without limb loss using three ADL-inspired tasks spanning gross motor manipulation, fine motor precision, and dynamic object interaction, informed by standardized functional assessments used in rehabilitation research^19–21^. Task-level performance metrics and subjective workload measures were used to quantify differences in efficiency, control, and perceived effort between end effectors^22^.

## Methods

### Participants

Twelve participants without limb loss (8 female, 4 male; age 25 *±* 3 years) were recruited for this study. All participants had prior experience using research-based myoelectric prosthetic systems, with a minimum of 120 minutes of cumulative exposure to the experimental prosthesis. To reduce learning effects while avoiding overtraining, participants had not used the device for at least six months prior to the study. Hand dominance was recorded (10 right-hand dominant, 2 left-hand dominant); however, the prosthesis was worn on the right arm for all trials. Although the HAMR framework is intended to support user-specific prosthetic design, a single HAMR-derived hand was used across all participants to establish a consistent baseline and isolate the effects of biologically informed geometry.

All participants provided informed consent before participation in the study. Experimental procedures were approved by the Johns Hopkins Medicine Institutional Review Board (IRB #00147458), and carried out in accordance with the Declaration of Helsinki and with the relevant guidelines and regulations set forth by the Johns Hopkins Medicine Institutional Review Board. Participants were compensated at a rate of $15 per hour, and the total experimental session lasted approximately 150 minutes.

### Experimental Apparatus

Experiments were conducted using the Tendon Actuated Modular Prosthesis (TAMP) (shown in Figure 1), a modular agonist/antagonist structured tendon-actuated prosthetic platform. TAMP consists of a forearm-mounted socket and shield assembly that obscures the participant’s biological hand and provides a standardized mounting interface for interchangeable prosthetic end effectors. End effectors were attached via a modified quick-disconnect wrist interface, enabling rapid, repeatable switching between conditions. Prosthetic actuation was driven by two rotary DC motors (Maxon RE30) mounted proximally within the TAMP structure. Each motor was equipped with an optical encoder (US Digital, 5000 counts per revolution) for position measurement and controlled via a current amplifier (Maxon ESCON 70/10). Surface electromyography (sEMG) signals were acquired from the wrist flexor and extensor muscle groups using an eight-channel sEMG system (Delsys Bagnoli)^18^.

**Figure 1.**
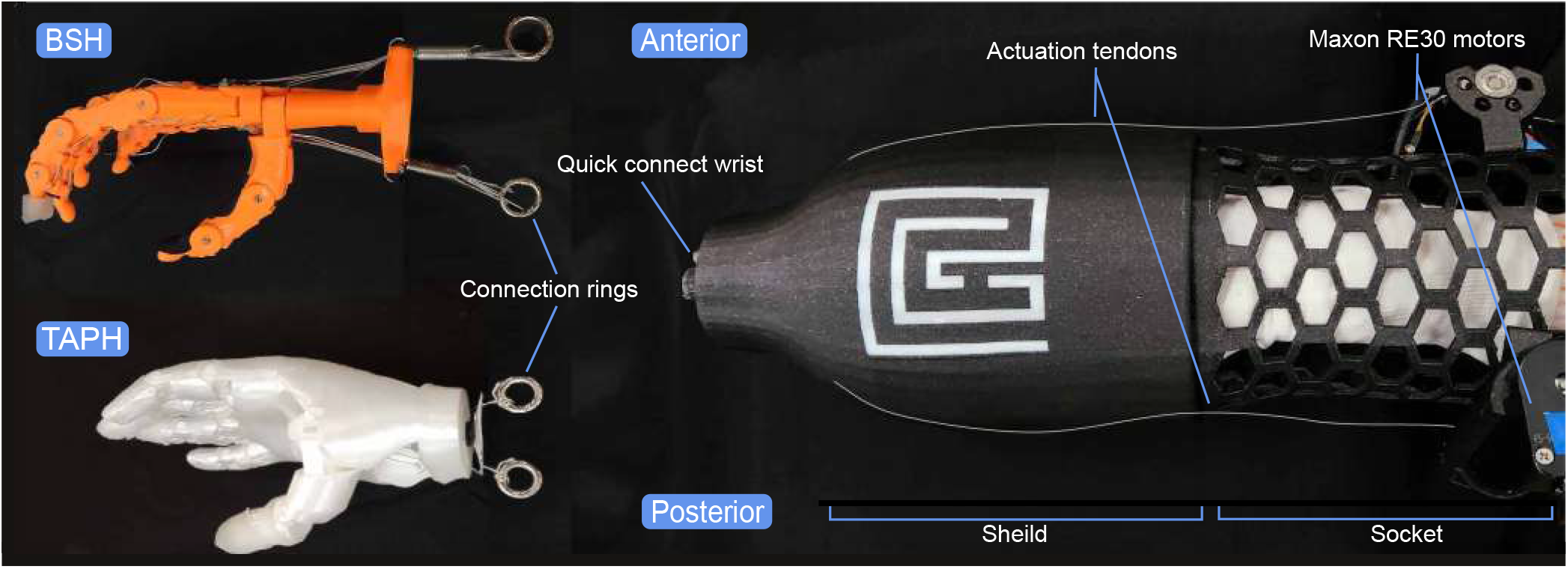
TAMP is a mock prosthesis featuring a shield with an integrated socket and a lattice with built-in mounts for the actuation motors. Tendons run from the motors to the connection rings of either the BSH or TAPH end effectors.

All sensing and actuation signals were processed in real time using a data acquisition system (Quanser Q8-USB) operating at a 1 kHz sampling rate. Control algorithms and data logging were implemented in MATLAB/Simulink (MathWorks) using QUARC real-time software. The same hardware and control configuration was used across all experimental conditions.

### Prosthetic End Effectors

Two tendon-driven prosthetic hand end effectors were evaluated in this study: the Bionic Skeletal Hand (BSH) and the Tendon Actuated Prosthetic Hand (TAPH) (shown in Figure 2). Both end effectors were designed for use with the Tendon Actuated Modular Prosthesis (TAMP) platform and were actuated using the same two-motor agonist/antagonist tendon configuration and identical control architecture. This design enabled controlled comparison between end effectors while holding tendon actuation, sensing, and control constant.

**Figure 2.**
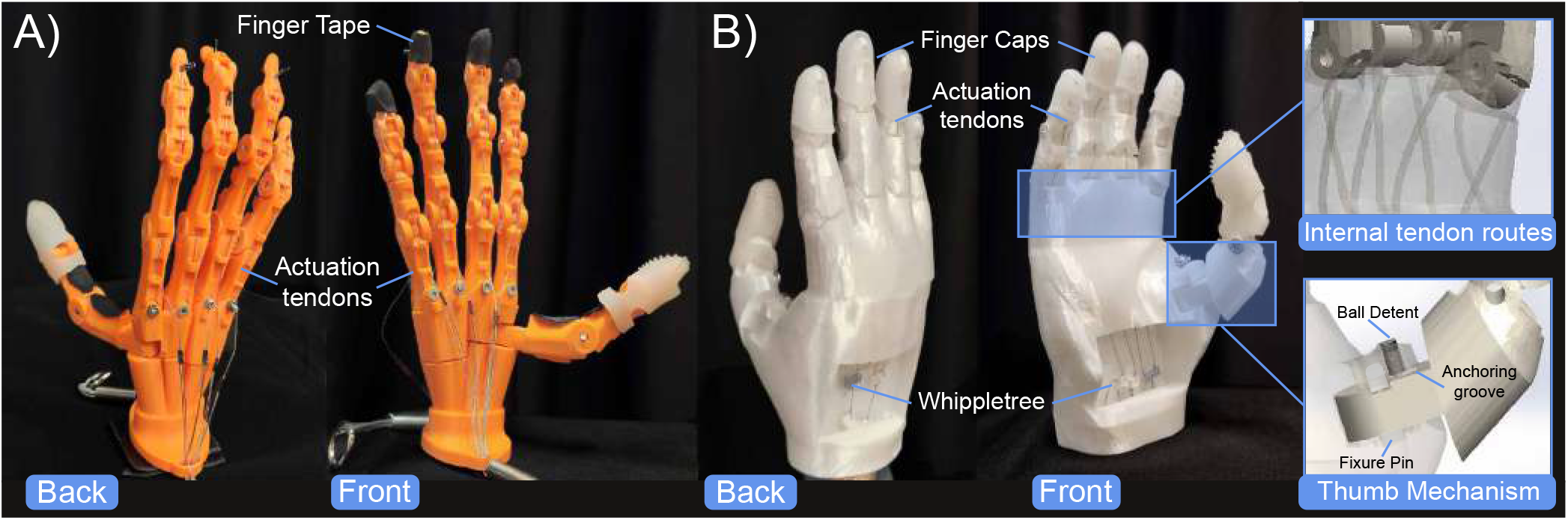
Internal tendon routing and structural components of (A) the BSH and (B) TAPH. TAPH includes a whippletree mechanism for even force distribution and features a semi-passive thumb for adaptable grasping.

**Figure 3.**
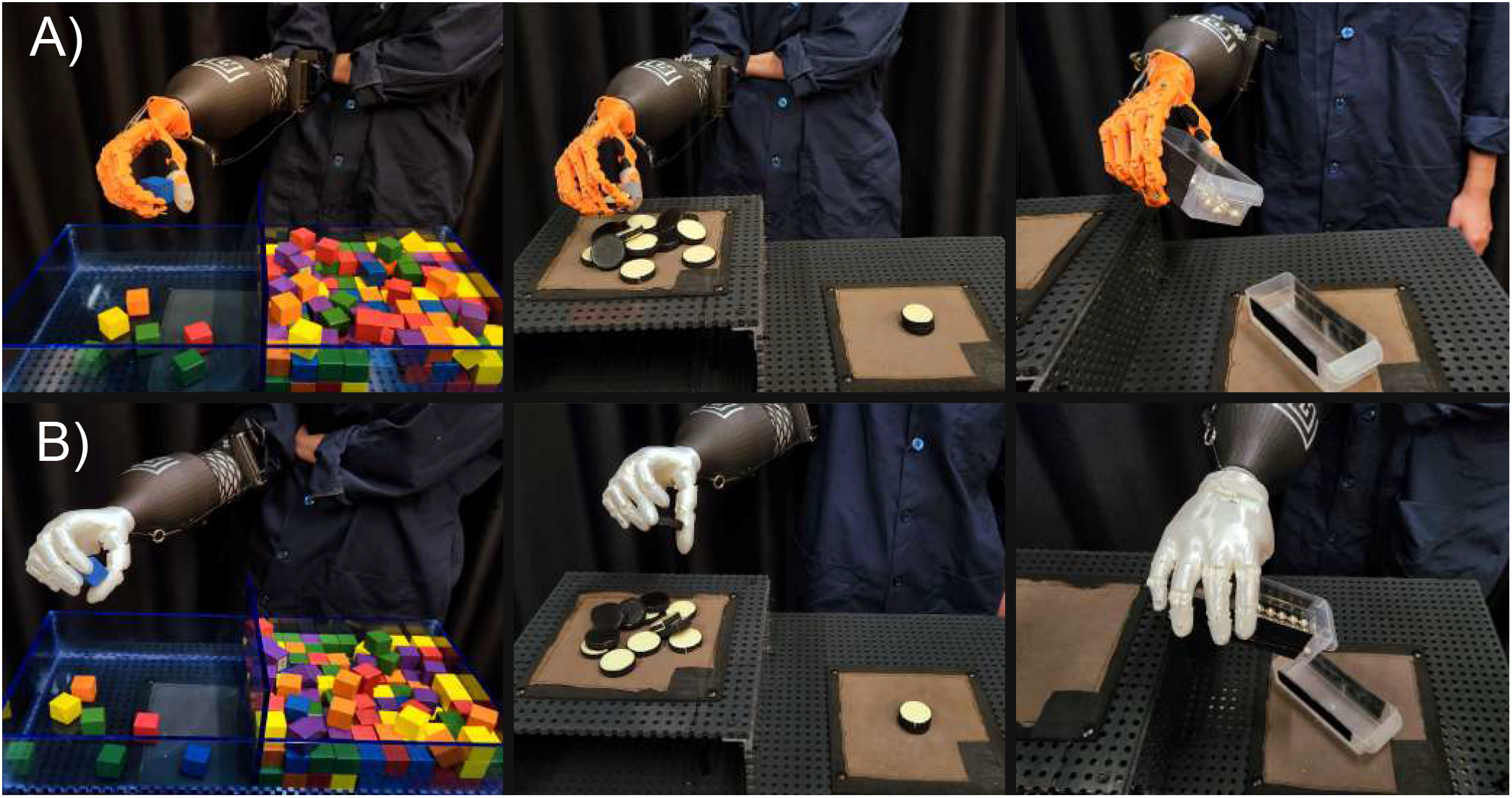
A participant performing the block transfer, checker transfer and stack, and carton transfer and pour tasks using (A) the BSH and (B) the TAPH end effectors.

#### Bionic Skeletal Hand (BSH)

The Bionic Skeletal Hand (BSH) is an anthropomorphic, tendon-driven prosthetic hand previously developed within our laboratory^13^. The hand comprises four articulated fingers, each with three actuated interphalangeal joints, resulting in a total of twelve actuated joints. The thumb is passively positioned and rotates as a single fused unit relative to the palm.

Flexion and extension tendons are routed along the anterior and posterior aspects of each finger, respectively, with tendons terminating at the distal end of the finger. Tendons from each finger converge at a common connection ring, which interfaces with the proximal actuation system of TAMP. This configuration enables simultaneous flexion and extension of all digits through the agonist/antagonist motor pair.

The BSH was fabricated using fused deposition modeling (FDM) additive manufacturing and assembled using rigid joint pins and tendon guides. The overall hand geometry reflects a generalized anthropomorphic form factor, with elongated finger segments and a minimized palmar region. Detailed depictions of BSH grasp types can be found in the Supplementary document.

#### Tendon Actuated Prosthetic Hand (TAPH)

The Tendon Actuated Prosthetic Hand (TAPH) was developed using the HAMR process to generate a geometry derived from human hand morphology (for details refer to supplementary document). TAPH includes four articulated fingers, each comprising two actuated interphalangeal joints, resulting in a total of eight actuated joints. The thumb is non-actuated and incorporates a semi-passive carpometacarpal joint that can be repositioned between lateral, pinch, and power grasp postures.The joint rotates about a fixed pin and transitions between grasp configurations via indexed grooves that engage a spring-loaded ball detent (see Figure 2).

As with BSH, flexion and extension tendons are routed along the anterior and posterior sides of each finger and driven by the same two-motor agonist/antagonist configuration. Tendons originating from the fingers converge proximally and interface with the TAMP actuation system. Internal tendon routing guides and joint alignments are defined based on the scanned hand geometry produced during the HAMR workflow.

TAPH was fabricated using FDM additive manufacturing and assembled using rigid joint pins and tendon routing features integrated into the printed structure. The hand geometry preserves human-derived finger proportions, joint-axis placement, and thumb orientation captured during the HAMR process. Detailed depictions of TAPH grasp types can be found in the Supplementary document.

### Force Distribution and Underactuation

Both prosthetic end effectors evaluated in this study are underactuated, having fewer actuators than kinematic degrees of freedom. BSH includes twelve actuated joints driven by two motors, while TAPH includes eight actuated joints driven by the same two-motor configuration. In both designs, flexion and extension tendons are routed through multiple joints within each finger, enabling joint-level adaptive motion during finger closure.

At the level of the full hand, the two end effectors differ in how actuation forces are distributed across fingers. In the BSH, individual finger tendons are rigidly coupled at a common connection ring driven directly by a motor-tendon unit. As a result, flexion and extension of all fingers occur simultaneously, and constraint of motion in any single finger limits further actuation of the remaining fingers, resulting in rigid underactuation.

To improve grasp conformity during object interaction, TAPH incorporates a force distribution mechanism that connects individual finger tendons to a Whippletree-based stage before routing to the actuation motors (see Figure 2). This mechanism equalizes tendon forces while allowing relative finger displacement. Consequently, if motion in one or more fingers is constrained by object contact, remaining fingers may continue to close and adapt to varying object geometry. This configuration results in adaptive underactuation at the hand level, while preserving identical actuator count, control inputs, and control strategy between end effectors. The inclusion of adaptive underactuation in TAPH improves grasp conformity and more closely resembles the actuation of the biological hand. Detailed depictions of TAPH grasp types can be found in the Supplementary document.

### Control and Calibration

The control scheme follows a First-In-First-Out Finite State Machine (FIFO FSM) control method to map sEMG signals to tendon actuation. Each participant undergoes an individualized calibration process to define flexor and extensor activation thresholds, ensuring robust signal differentiation. The control scheme allows for simultaneous tendon activation, meaning that if co-contraction occurs, the system actuates both tendons accordingly. These activation thresholds help ensure clarity of movement, along with a defined idle state, thereby creating the four control states (Flexion, Extension, Co-contraction, and Neutral) as shown in Table S1 of the Supplementary document. To control the prosthesis in the Flexion, Extension, and Co-contraction actuation states, the sEMG signal is proportionally mapped to the respective motor current as originally detailed in^13^ and as shown in:

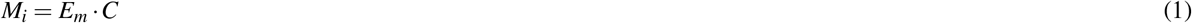

Where *M*_*i*_ is the motor command for motor *i* (either agonist or antagonist), *E*_*m*_ represents the sEMG input for either muscle group *m*, and the constant *C* represents a participant-specific scaling and conversion factor that takes each participant’s EMG calibration into account. The primary motor during actuation refers to the motor aligned with the intended direction of actuation (e.g., during flexion, the anterior motor is the primary motor) and rotates clockwise to actuate the hand. At the same time, the secondary motor, or opposing motor in the movement, proportionally spins counterclockwise to facilitate movement. This collaborative movement enables smooth actuation of the prosthesis. In the co-contraction case, the motors both pull in the clockwise direction.

### Experimental Tasks and Procedure

#### Experimental Procedure

At the start of each session, participants were briefed on the experimental procedure and trained on the control motions used to operate the prosthesis: wrist flexion to activate the flexor muscle group, wrist extension to activate the extensor muscle group, and relaxation to return to a neutral state. Following electrode placement and sEMG calibration, participants were assisted in donning the prosthesis and given up to two minutes to practice with a simplified version of each task to familiarize themselves with the control scheme.

Each participant completed three trials of three experimental tasks for a total of nine trials. Each trial lasted two minutes. The order of tasks and prosthetic hand conditions (BSH or TAPH) was randomized and counterbalanced across participants. An automated auditory cue signaled the start of each trial, the midpoint, and the conclusion of the trial. A two-minute rest period was provided between trials, during which participants completed a NASA Task Load Index (TLX) questionnaire.

All trials were performed while participants stood at a standardized workstation. Wrist orientation was fixed relative to the workspace and remained constant across all trials. Likewise, thumb position remained fixed in the pinch orientation for both end effectors and was not changed between tasks.

#### Experimental Tasks

Three ADL-inspired tasks were used to evaluate prosthetic performance across gross motor manipulation, fine motor precision, and dynamic object interaction. Task design was informed by standardized functional assessments commonly used in rehabilitation research^19–23^.

##### Box and Blocks Task

Participants performed a modified Box and Blocks task in which they transferred wooden blocks (each measuring 2.5 cm^3^) from one compartment of a divided box to the other, one block at a time, using the prosthetic hand. The task required repeated grasping, lifting, and placement of blocks within the two-minute trial period. The total number of blocks transferred was recorded for each trial.

##### Checker Transfer and Stack Task

In the checker transfer and stack task, participants grasped individual checkers (3.8 cm in diameter and 1 cm thick) from an elevated surface and transferred them to a lower surface (11 cm vertical difference), stacking the checkers as they were placed. This task required precise grasp formation and placement to successfully stack thin, flat objects. The total number of checkers transferred and the number of checkers successfully stacked were recorded for each trial.

##### Carton Transfer and Pour Task

The carton transfer and pour task required participants to grasp a rectangular carton (5 cm wide, 15 cm long, 5 cm tall) containing weighted marbles (50, 75, 100g), transfer it between two surfaces of different heights (11 cm), and pour the contents into an identical empty carton. Participants repeated this sequence as many times as possible within the trial period. This task introduced dynamic changes in object mass distribution during manipulation. The number of successful transfers, the number of pours, and pour accuracy were recorded for each trial.

### Metrics

The following metrics were used to analyze participants’ performance across trials.

#### # of Blocks Transferred

The number of blocks transferred indicates the total quantity of blocks that a participant moves from one side of the box to the other within a trial of the box and blocks task.

#### # of Checkers Transferred

The number of checkers transferred represents the total quantity of checkers that a participant successfully moves from one side of the transfer table to the other during a given trial of the checker transfer and stack task.

#### # of Checkers Stacked

The number of checkers stacked refers to the total number of checkers that a participant successfully stacks on top of one another during a trial of the checker transfer and stack task.

#### # of Carton Transfers

The number of carton transfers refers to the total count of successful transfers completed during a trial of the carton transfer and pour task. A successful transfer is defined as the uninterrupted movement of a carton from one side of the transfer table to the other.

#### # of Carton Pours

The number of carton pours is defined as the total instances in which a participant successfully pours the weighted marbles from one carton into the other during a trial of the carton transfer and pour task.

#### Carton Pour Accuracy

Carton pour accuracy is determined by calculating the proportion of marbles successfully transferred during each pour relative to the total marble count, producing an average accuracy score for each trial of the carton transfer and pour task.

### Statistical Analysis

All statistical analyses were conducted using RStudio (v2.1) to assess participant performance on each task. Prior to conducting parametric tests, we confirmed that the data met the assumptions of normality and homogeneity of variance. A random intercept linear mixed effects model was applied to analyze key metrics for each task and survey rating results. In each model, participants were treated as random effects, while end effector type was treated as a fixed effect. For the carton transfer task, weight was included as an additional fixed effect. Continuous variables in the analysis included total transfers for each task, checker stacks, and carton pours. Trial number was analyzed as a covariate in each model to assess learning effects and was treated as a continuous variable. To account for multiple comparisons, a Tukey correction was applied, with significance set at a p-value threshold of 0.05.

## Results

Task performance and subjective workload differed between prosthetic end effector conditions across all experimental tasks. Linear mixed-effects models were used to assess the effects of end effector condition and task-specific factors on performance outcomes. Trial number was used to evaluate the impact of natural learning in the task.

### Block Transfers Task

Participants completed a greater number of block transfers when using TAPH compared to BSH (*β* = −10.08, SE = 1.08, p < 0.001). Across both conditions, trial number did not have a significant effect on block transfer performance (*β* = −0.32, SE = 0.26, p = 0.22). Group-level performance across trials is shown in Figure 4.

**Figure 4.**
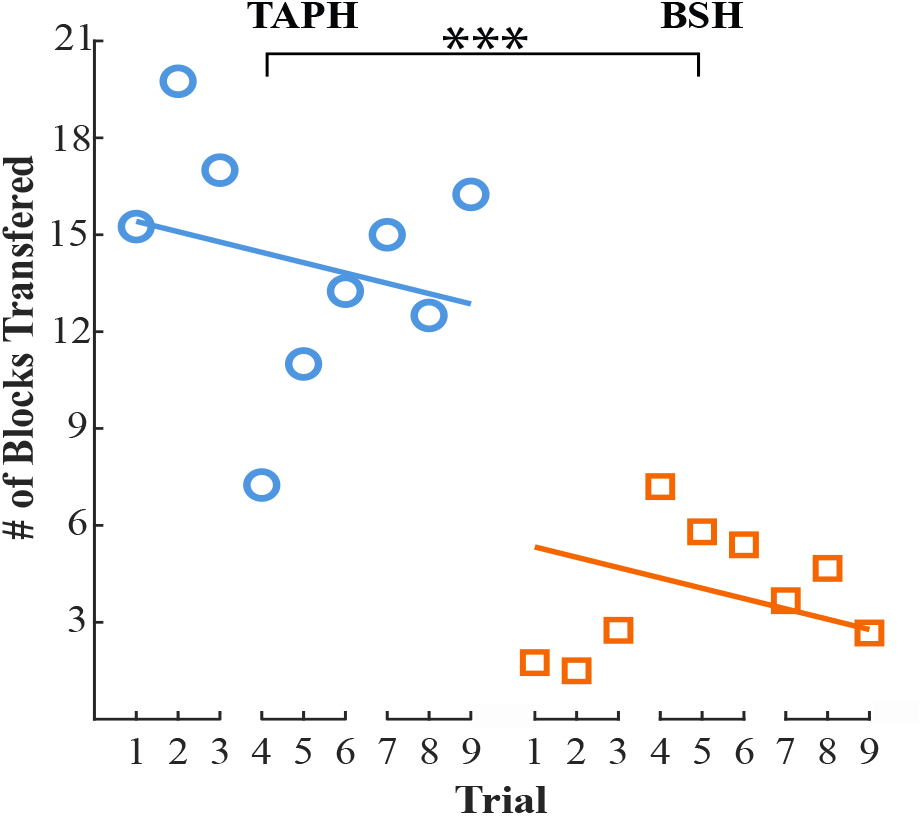
The average number of block transfers for each condition across trials. Individual data points represent the average for each trial (for all participants), and the solid lines indicate the model’s predictions. Trials 1-3 represent results from participants who completed the block transfer task first, trials 4-6 correspond to those who completed the task second, and trials 7-9 show results for participants who completed the task third. * indicates p < 0.05, ** indicates p < 0.01, and *** indicates p < 0.001.

### Checker Transfer and Stack Task

For the checker transfer task, participants transferred more checkers when using TAPH than when using BSH (*β* = −3.08, SE = 0.56, p < 0.001). Trial number did not have a significant effect on checker transfer performance (*β* = −0.20, SE = 0.147, p = 0.16). For the checker stacking task, the number of checkers successfully stacked was higher when using TAPH compared to BSH (*β* = −3.72, SE = 0.51, p < 0.001). Trial number had a significant effect on stacking performance across conditions (*β* = 0.30, SE = 0.13, p = 0.02). Performance trends for checker transfer and stacking are shown in Figure 5.

**Figure 5.**
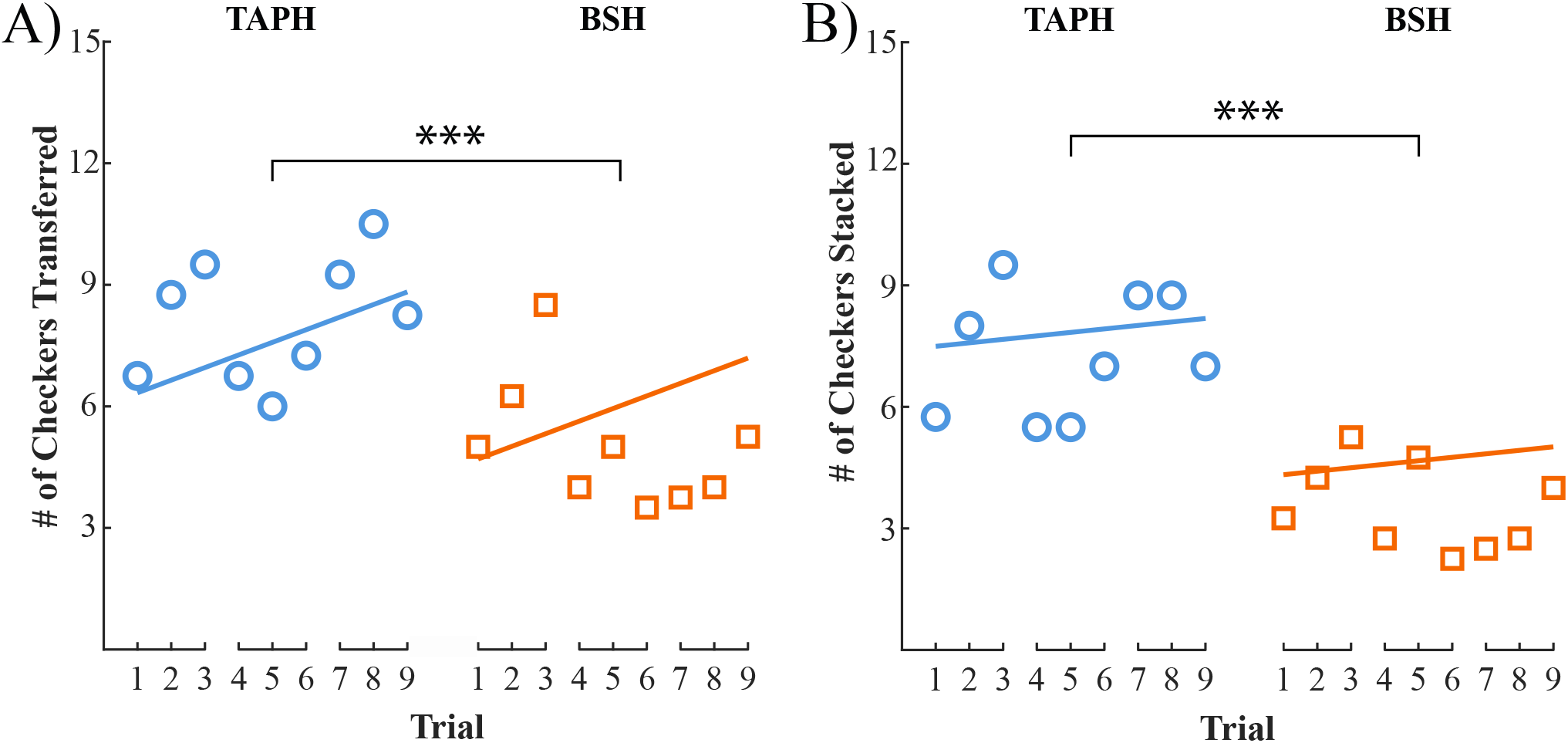
The average number of (A) checker transfers, and (B) checkers stacked for each condition across trials. Where the individual data points represent the average for each trial (for all participants), and the solid lines indicate the model’s predictions. Trials 1-3 represent results from participants who completed the block transfer task first, trials 4-6 correspond to those who completed the task second, and trials 7-9 show results for participants who completed the task third. * indicates p < 0.05, ** indicates p < 0.01, and *** indicates p < 0.001.

### Carton Transfer and Pour Task

Participants completed a greater number of carton transfers when using TAPH compared to BSH (*β* = −1.63, SE = 0.57, p < 0.01). Both trial number (*β* = 0.29, SE = 0.145, p = 0.04) and carton weight (*β* = −1.57, SE = 0.35, p < 0.001) had significant effects on the number of carton transfers completed. Similarly, the number of carton pours was higher when using TAPH than BSH (*β* = −1.63, SE = 0.57, p < 0.01). Carton weight had a significant negative effect on pour count (*β* = −1.47, SE = 0.35, p < 0.001), while trial number did not reach significance (*β* = 0.26, SE = 0.147, p = 0.07). Carton transfer and pour performance is shown in Figure 6.

**Figure 6.**
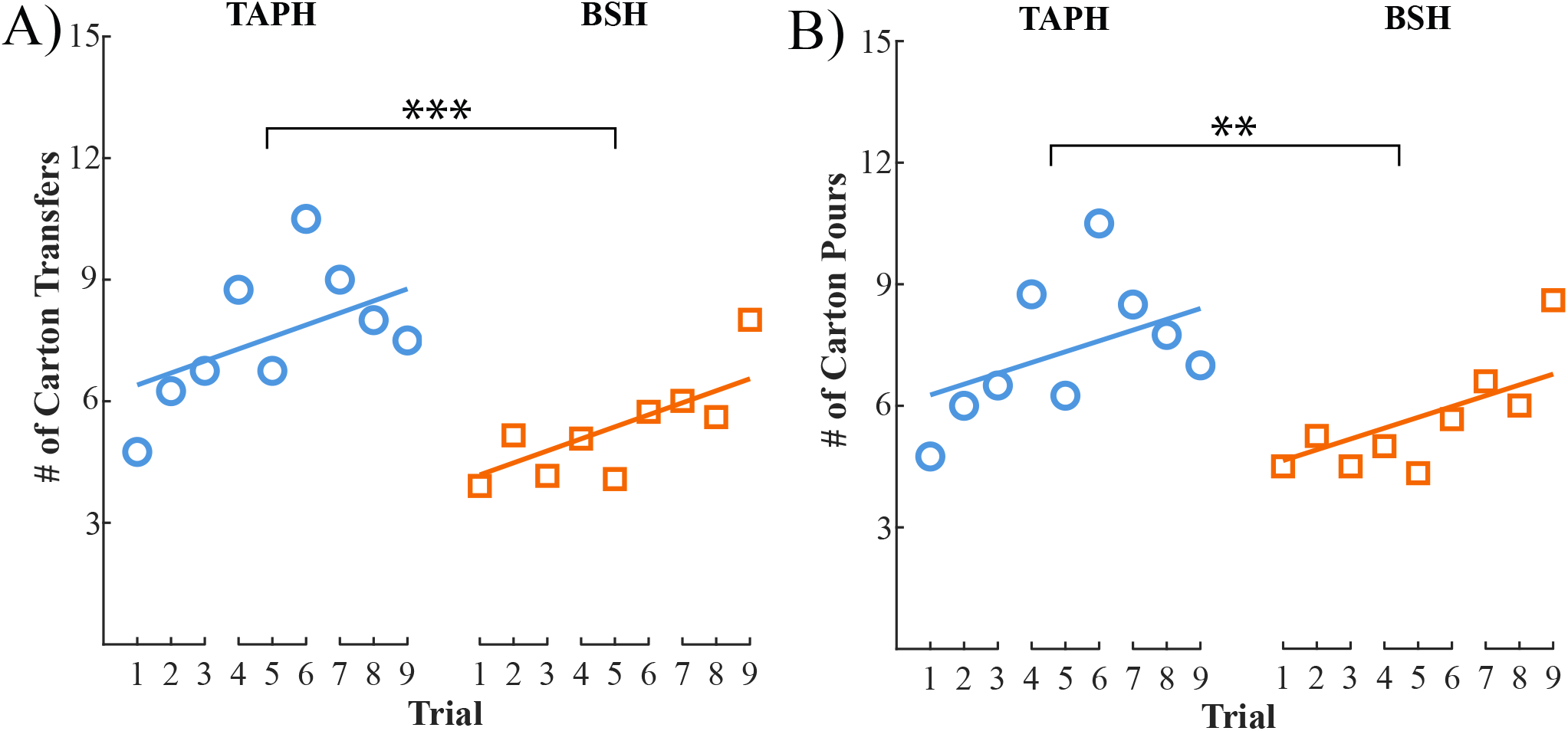
The average number of (A) carton transfers and (B) carton pours for each condition across trials. Individual data points represent the average for each trial (for all participants), and the solid lines indicate the model’s predictions. Trials 1-3 represent results from participants who completed the block transfer task first, trials 4-6 correspond to those who completed the task second, and trials 7-9 show results for participants who completed the task third. * indicates p < 0.05, ** indicates p < 0.01, and *** indicates p < 0.001.

### Carton Pour Accuracy

Carton pour accuracy did not differ significantly between end effector conditions (*β* = 0.005, SE = 0.04, p = 0.90). Across both conditions, trial number had a significant effect on pour accuracy (*β* = 0.02, SE = 0.009, p = 0.01), while increased carton weight was associated with reduced accuracy (*β* = −0.10, SE = 0.02, p < 0.001). Accuracy trends are shown in Figure 7.

**Figure 7.**
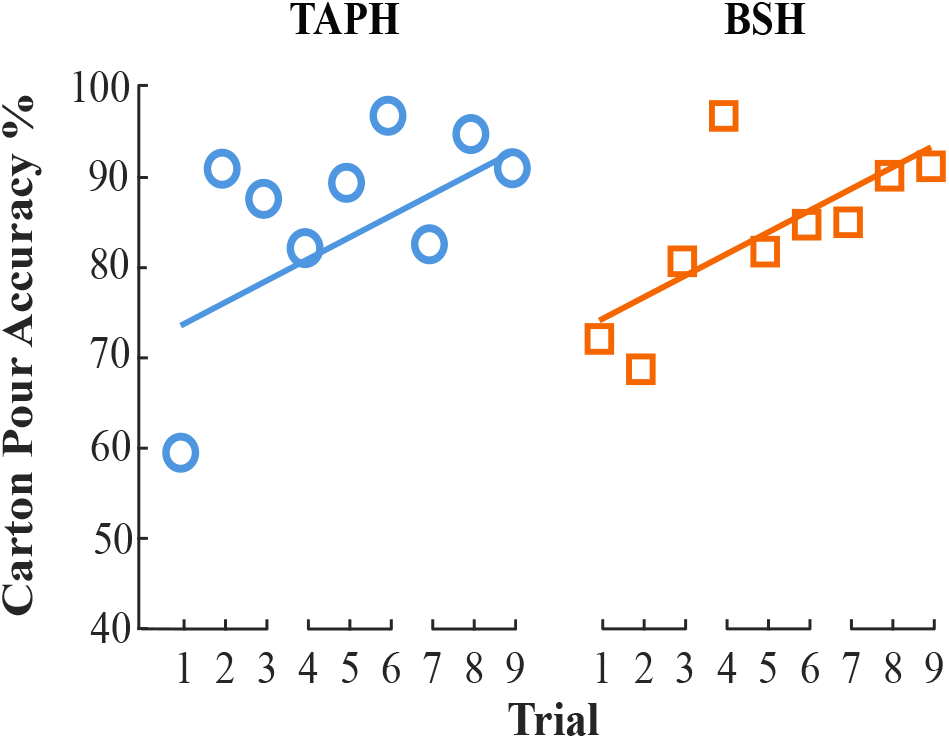
The average carton pour accuracy for each condition across trials. Where the individual data points represent the average for each trial (for all participants), and the solid lines indicate the model’s predictions. Trials 1-3 represent results from participants who completed the block transfer task first, trials 4-6 correspond to those who completed the task second, and trials 7-9 show results for participants who completed the task third. * indicates p < 0.05, ** indicates p < 0.01, and *** indicates p < 0.001.

### Subjective Workload: NASA TLX

Subjective workload ratings differed between end effector conditions across tasks (Figure 8). During the block transfer task, participants reported lower mental demand, physical demand, effort, and frustration, as well as higher perceived performance when using TAPH compared to BSH (all p < 0.001). Similar patterns were observed during the checker transfer and stacking task, with lower mental demand, physical demand, effort, and frustration, and higher perceived performance for TAPH (all p < 0.001). For the carton transfer and pour task, effort (p < 0.05) and frustration (p < 0.005) ratings were lower when using TAPH, while no significant differences were observed in mental demand, physical demand, or perceived performance between end effectors.

**Figure 8.**
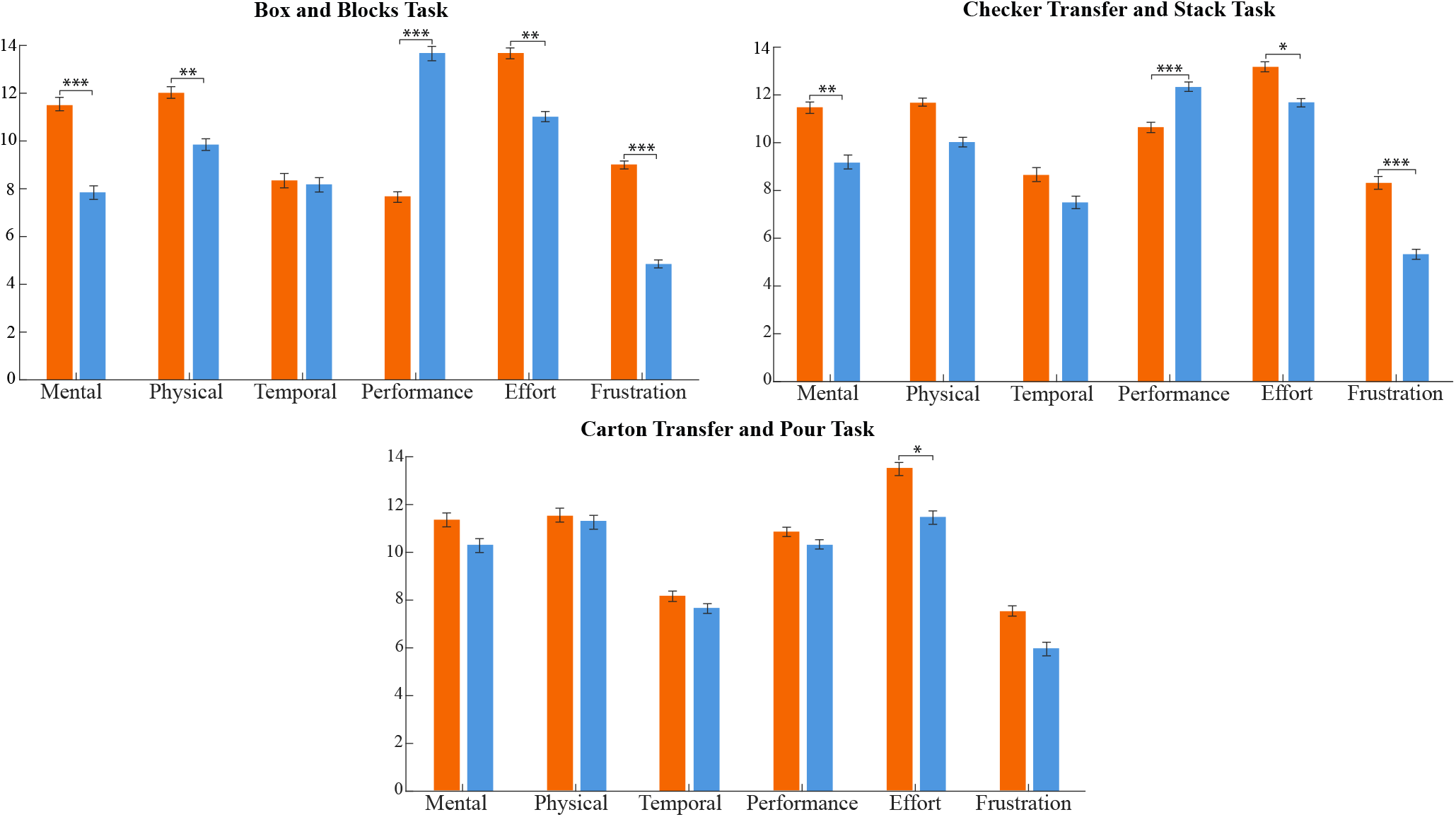
TLX workload ratings for the block transfer (top left), checker transfer (top right), and carton transfer (bottom) tasks across end effector conditions. Each bar represents the mean subjective score across participants for each subscale: mental demand, physical demand, temporal demand, performance, effort, and frustration. * indicates p < 0.05, ** indicates p < 0.01, and *** indicates p < 0.001.

## Discussion

This study evaluated the functional impact of biologically informed prosthetic hand design through a controlled comparison of two tendon-driven end effectors that differed in geometric mapping and force distribution strategy while sharing identical actuation and control architectures. Across a range of ADL tasks, use of the Tendon Actuated Prosthetic Hand (TAPH) was associated with higher task performance and lower subjective workload relative to a generalized Bionic Skeletal Hand (BSH). These results indicate that biologically driven hand design features play a meaningful role in functional task outcomes during prosthetic use, even when actuation and control are held constant.

In the block transfer task, TAPH enabled a higher rate of object transfers than the BSH, with no significant effect of trial number across conditions. The absence of a learning effect suggests that observed differences were driven by intrinsic properties of the end effector designs rather than short-term familiarization. Gross motor manipulation emphasizes repeated grasp formation and release under relatively stable contact conditions, placing greater demands on grasp robustness and consistency than on fine positional accuracy. The improved performance observed with TAPH in this context is consistent with design features that support stable multi-digit contact and efficient force transmission during grasping.

Performance differences persisted in the checker transfer and stacking task, which required greater precision during grasp formation and object placement. TAPH was associated with higher transfer and stacking counts, while a trial effect was observed only for stacking performance, suggesting that learning was more strongly associated with fine object placement. These results suggest that biologically informed hand geometry and force distribution can enhance fine motor performance under stable interaction conditions, particularly when precision actions are repeatedly required.

In contrast, the dynamic carton transfer and pour task revealed more nuanced effects. While participants completed a greater number of transfers and pours using TAPH, no significant difference in pour accuracy was observed between end effectors. This task introduced dynamic changes in object mass distribution, increasing demands on continuous coordination during interaction. The convergence in accuracy outcomes suggests that advantages observed under stable conditions may not fully generalize to tasks requiring ongoing adjustment in response to changing object dynamics, particularly when employing prosthetic devices that lack an actively controlled wrist degree of freedom, resulting in a need for frequent repositioning and constraining performance during pouring. These outcomes show that anthropomorphic hand geometry and force distribution alone are insufficient to fully support dynamic fine motor tasks that require continuous reorientation and adjustment and highlight the task-dependent nature of prosthetic performance.

Although both end effectors evaluated in this study are underactuated and controlled using identical motor and control architectures, they differ in how actuation forces are distributed across the hand. At the finger level, both designs permit joint-level adaptive motion through multi-joint tendon routing. At the hand level, however, TAPH incorporates a Whippletree-based force distribution mechanism that enables adaptive underactuation across digits, allowing individual fingers to continue closing when others are constrained by object contact (similar to a differential in a geared system). By contrast, the rigid coupling of finger tendons in the BSH limits further motion when any digit encounters resistance. Importantly, adaptive underactuation was an intentional design feature of TAPH, motivated by biological grasping behavior in which force redistribution across digits supports more natural and robust interaction with irregular objects. The improved performance observed with TAPH in gross motor and stable fine motor tasks is therefore consistent with the combined influence of biologically informed geometry and adaptive force distribution, rather than differences in actuator count or control strategy.

In addition to force distribution, task performance was influenced by differences in the kinematic structure and anatomical mapping of the two end effectors. The improved performance observed with TAPH can be directly attributed to multiple HAMR-derived geometric features that shape grasp kinematics. The anthropometric placement and spacing of the TAPH thumb enabled participants to employ grasp strategies that more closely resembled natural hand use compared to the BSH. In particular, the thumb could directly oppose the index and middle fingers to stabilize objects during transfer and pouring tasks, or to manipulate slender objects during checker handling, while the compliant interphalangeal joint maintained surface contact during grasp closure.

Although both devices employed passively adjustable thumbs, the orientation of the BSH thumb often required participants to curl the fingers into the palm to establish object contact, which limited effective manipulation in tasks requiring opposition-based grasping. In addition, palm to finger proportions, thumb orientation, and joint axis placement in the BSH diverged from human hand anatomy. By contrast, TAPH preserved metacarpophalangeal joint orientation, palm to finger proportions, and thumb opposition geometry directly mapped from a human hand scan. These kinematic differences provide a plausible explanation for how biologically mapped geometry enabled adaptive mechanics to be expressed through more natural and intuitive grasp strategies during task execution.

Subjective workload ratings further substantiated the observed task performance differences between the two end effectors across tasks. Use of TAPH was associated with lower perceived mental demand, physical demand, effort, and frustration, particularly in tasks involving repeated or precise grasping. These reductions suggest that biologically informed design and adaptive force distribution may reduce the cognitive and physical effort required to achieve task goals. In the dynamic carton transfer and pour task, workload differences were more limited, as participants were required to manage dynamic changes in object weight and orientation, which increased the need for sustained attention and adjustment during manipulation. As a result, users likely relied more heavily on compensatory strategies to maintain control and achieve task goals. This increased attentional demand provides a plausible explanation for why perceived cognitive workload converged between end effectors, despite differences in task performance.

Together, these findings demonstrate that biologically informed hand design can meaningfully influence prosthetic performance and user experience, particularly in tasks involving stable object interaction. While increased anthropomorphic complexity alone does not guarantee functional improvement, the present results indicate that targeted incorporation of biological design principles, such as anatomically mapped geometry and adaptive underactuation, can enhance task efficiency without increasing complexity in device control. Future work should examine how these design features interact with individualized morphology, sensory feedback, and adaptive control strategies, particularly in users with limb loss and during extended real-world use.

## Data Availability

The datasets used and/or analyzed during the current study are available from the corresponding author on reasonable request.

## Acknowledgements

L.V. Author thanks Chase Lahr and Emmaleigh Shinno for their work on the manufacturing of the hand device and the National Science Foundation (grant# 2146206) for funding this research.

## Author contributions statement

L.V. and J.D.B. conceived the experiment, L.V. conducted the experiment, and L.V. analyzed the results. All authors reviewed the manuscript.

## Declarations

### 0.0.1 Ethics approval

All participants provided informed consent before participation in the study. Experimental procedures were approved by the Johns Hopkins Medicine Institutional Review Board (IRB #00147458).

### 0.0.2 Consent for publications

All participants provided informed consent for publication as outlined by their participant consent and authorization form approved by the Johns Hopkins Medicine Institutional Review Board (IRB #00147458).

## References

1. Basumatary, H. & Hazarika, S. M. State of the art in bionic hands. IEEE Transactions on Human-Machine Syst.50, DOI: 10.1109/THMS.2020.2970740 (2020).

2. Huang, H. H., Hargrove, L. J., Ortiz-Catalan, M. & Sensinger, J. W. Integrating Upper-Limb Prostheses with the Human Body: Technology Advances, Readiness, and Roles in Human–Prosthesis Interaction. Annu. Rev. Biomed. Eng.26, 503–528, DOI:10.1146/annurev-bioeng-110222-095816 (2024).

3. Salminger, S. et al. Current rates of prosthetic usage in upper-limb amputees–have innovations had an impact on device acceptance?Disabil. Rehabil.44, DOI:10.1080/09638288.2020.1866684 (2022).

4. Spiers, A. J., Cochran, J., Resnik, L. & Dollar, A. M. Quantifying Prosthetic and Intact Limb Use in Upper Limb Amputees via Egocentric Video: An Unsupervised, At-Home Study. IEEE Transactions on Med. Robotics Bionics3, DOI: 10.1109/TMRB.2021.3072253 (2021).

5. Biddiss, E. & Chau, T. Upper limb prosthesis use and abandonment: A survey of the last 25 years, DOI:10.1080/03093640600994581 (2007).

6. Young, B. H. The Bionic-Hand Arms Race: High-Tech Hands are Complicated, Costly, and Often Impractical. IEEE Spectr.59, DOI:10.1109/MSPEC.2022.9915629 (2022).

7. Brown, J. D. et al. Touching reality: Bridging the user-researcher divide in upper-limb prosthetics. Sci. Robotics8, DOI: 10.1126/SCIROBOTICS.ADK9421 (2023).

8. Chappell, D. et al. Beyond Humanoid Prosthetic Hands: Modular Terminal Devices That Improve User Performance. IEEE Transactions on Neural Syst. Rehabil. Eng. (Early Access)1–1 (2025).

9. Correia, T., Ribeiro, F. M. & Pinto, V. H. Realistic Model Parameter Optimization: Shadow Robot Dexterous Hand Use-Case. In Communications in Computer and Information Science, vol. 1982 CCIS, DOI:10.1007/978-3-031-53036-4_17 (2024).

10. Fukaya, N., Toyama, S., Asfour, T. & Dillmann, R. Design of the TUAT/Karlsruhe humanoid hand. IEEE Int. Conf. on Intell. Robots Syst.3, DOI:10.1109/IROS.2000.895225 (2000).

11. Laffranchi, M. et al. The Hannes hand prosthesis replicates the key biological properties of the human hand. Sci. Robotics 5, DOI:10.1126/SCIROBOTICS.ABB0467 (2020).

12. Ajoudani, A. et al. Exploring teleimpedance and tactile feedback for intuitive control of the pisa/IIT soft hand. IEEE Transactions on Haptics7, DOI:10.1109/TOH.2014.2309142 (2014).

13. Miller, E., Amanze, I. & Brown, J. A Wearable Anthropomorphically-Driven Prosthesis with a Built-In Haptic Feedback System. InIn Proceedings of 2020 International Symposium on Medical Robotics, ISMR 2020, DOI:10.1109/ISMR48331.2020.9312933 (2020).

14. Belter, J. T., Leddy, M. T., Gemmell, K. D. & Dollar, A. M. Comparative clinical evaluation of the Yale Multigrasp Hand. InProceedings of the IEEE RAS and EMBS International Conference on Biomedical Robotics and Biomechatronics, vol. 2016-July, DOI:10.1109/BIOROB.2016.7523680 (2016).

15. Godfrey, S. B. et al. The Softhand Pro: Functional evaluation of a novel, flexible, and robust myoelectric prosthesis. PLoS ONE13, DOI:10.1371/journal.pone.0205653 (2018).

16. Cabibihan, J. J. et al. Suitability of the Openly Accessible 3D Printed Prosthetic Hands for War-Wounded Children. Front. Robotics AI7, DOI:10.3389/frobt.2020.594196 (2021).

17. Cempini, M., Cortese, M. & Vitiello, N. A powered finger-thumb wearable hand exoskeleton with self-aligning joint axes. IEEE/ASME Transactions on Mechatronics20, DOI:10.1109/TMECH.2014.2315528 (2015).

18. Velásquez, L. & Brown, J. D. Understanding the Utility of State-Based Haptic Feedback in Tendon-driven Anthropomorphic Prostheses. IEEE Transactions on Neural Syst. Rehabil. Eng.DOI:10.1101/2025.02.27.25323021 (2025).

19. Mathiowetz, V., Volland, G., Kashman, N. & Weber, K. Adult norms for the Box and Block Test of manual dexterity. The Am. journal occupational therapy. : official publication Am. Occup. Ther. Assoc.39, DOI:10.5014/ajot.39.6.386 (1985).

20. Sigirtmac, I. & Oksuz, C. Investigation of reliability, validity, and cutoff value of the Jebsen-Taylor Hand Function Test. J. Hand Ther.34, DOI:10.1016/j.jht.2020.01.004 (2021).

21. Vasluian, E. et al. Learning effects of repetitive administration of the southampton hand assessment procedure in novice prosthetic users. J. Rehabil. Medicine46, DOI:10.2340/16501977-1827 (2014).

22. Hart, S. G. & Staveland, L. E. Development of NASA-TLX (Task Load Index): Results of Empirical and Theoretical Research. Adv. Psychol.52, DOI:10.1016/S0166-4115(08)62386-9 (1988).

23. Thomas, N., Miller, A. J., Ayaz, H. & Brown, J. D. Haptic shared control improves neural efficiency during myoelectric prosthesis use. Sci. Reports13, DOI:10.1038/s41598-022-26673-2 (2023).

