## Supplemental Document for "Biologically informed geometry and force distribution improve task performance in agonist/antagonist tendon-driven prosthetic hands"

for

### Supplementary Information

#### TAPH and BSH Grasp Types

To illustrate the functional grasp strategies enabled by each device, Figure S2 presents representative grasp postures achieved with the BSH and TAPH, respectively. Both devices support power, pinch, and lateral grasps, but the underlying mechanics differ.

In BSH, grasping relies on curling elongated fingers into a minimized palm, with a passively oriented thumb that offers limited opposition. This configuration supports gross grasps but often compromises precision and stability. By contrast, TAPH preserves human-like palmar proportions and joint axis orientations, while incorporating an anthropomorphic thumb design. The thumb has two key features: (1) a carpometacarpal (CMC) detent mechanism that locks into three discrete postures: lateral, pinch/tripod, and power, and (2) a passive interphalangeal (IP) joint that flexes compliantly to conform to object surfaces. Together, these features enable more natural pinch and tripod grasps, stable power grasps, and improved contact with slender or irregular objects (e.g., checkers). The compliant IP joint prevents collapse into full extension by maintaining surface contact against the object, distributing forces more evenly across the grasp. These differences highlight how anthropomorphically mapped geometry and compliant thumb mechanics in TAPH provide functional benefits beyond simple joint count, improving precision and reducing compensatory wrist and shoulder strategies compared to BSH.

#### Human-inspired Actuator Modeling and Reconstruction (HAMR) Process

The Human-Inspired Actuator Modeling and Reconstruction (HAMR) process is a design framework for generating anatomically informed, tendon-actuated prosthetic hand geometric implements. Rather than prescribing a fixed device geometry, the HAMR process leverages subject-specific anatomical information to guide the placement of joints, tendon routing channels, and thumb mechanisms.

In the present work, the HAMR process was demonstrated using a mold of a human hand captured in a relaxed, open posture. The resulting cast was digitized using a handheld 3D scanner (Revopoint Pop 3) to generate a surface representation of the hand geometry. This digital model was converted into an editable solid model and served as the basis for incorporating prosthesis-specific design features, including articulated finger joints, internal tendon routing channels, and structural cavities for differential force distribution via a Whippletree mechanism. A semi-fixed thumb joint was incorporated to support lateral, pinch, and power grasp configurations, reflecting common functional thumb postures during activities of daily living.

The finalized geometry was fabricated using additive manufacturing and assembled with routed tendons to produce a functional tendon-actuated prosthetic hand. Figure S2 illustrates the overall progression of the HAMR workflow from physical casting to the assembled, 3D-printed device. In the current study, a single hand geometry was used across participants to isolate the effects of end effector design on task performance. Ongoing and preliminary work extending the HAMR process toward individually manufactured end effectors and partial-hand applications is discussed below.

### The HAMR Process: Partial Hand

To demonstrate the versatility of the HAMR process beyond full hand replacements, we initiated a pilot study extending the framework to design a custom prosthetic fitting for a partial hand amputee. The user, who regularly used individual finger prostheses from Naked Prosthetics, experienced difficulty maintaining consistent use due to the length and geometry of her residual digits, which often prevented secure, comfortable fitting. She expressed interest in two key adaptations: (1) custom flexible fittings to improve anchoring of her residual fingers to the Naked Prosthetics devices, and (2) the development of a tendon-driven, agonist/antagonist partial hand design that could integrate with these fittings. We employed the HAMR Process by first capturing the geometry of the user's residual limb through creating a stone plaster mold. We then used a Revopoint Pop 3 3D scanner to generate an STL. A multi-material design approach was employed to fabricate a flexible, press-fit base that conformed to the residual anatomy and could be integrated into either the existing Naked Prosthetic fingers or rigid tendon-actuated finger modules to restore grasping function (see Figure S2). The resulting design (shown in Figure S3) maintains anatomical alignment while enabling active tendon-driven motion of the prosthetic fingers. This case highlights the potential of the HAMR process to generate low-cost, anatomically matched solutions for individuals with partial limb loss. Future work will build on this pilot by refining the multi-material interface, expanding control strategies for partial hand actuation, and evaluating the HAMR process across a broader range of partial hand presentations to support fully customized, functionally integrated prosthetic solutions.

### Supplementary Tables

Table S1: TAMP Control States and Threshold Conditions

| State | Condition | Direction | |
|---|---|---|---|
| | | Antagonist Motor | Posterior Motor |
| Flexion | Ant_EMG > $Ant_{th}$ | $Ant_m$ = CW | $Post_m$ = CCW |
| Extension | Post_EMG > $Post_{th}$ | $Ant_m$ = CW | $Post_m$ = CCW |
| Cocontraction | Ant_EMG > $Ant_{th}$ and Post_EMG > $Post_{th}$ | $Ant_m$ = CW | $Post_m$ = CW |
| Neutral | Otherwise | $Ant_m = 0$ | $Post_m = 0$ |

Control logic used to determine state transitions based on EMG signals from the flexor (Ant_EMG) and extensor (Post_EMG) muscle groups. Each row defines a distinct control state and the corresponding threshold-based condition required to activate that state. Threshold values are denoted by $Ant_{th}$ and $Post_{th}$, and motor outputs are denoted as $Ant_m$ and $Post_m$, with clockwise (CW) and counter-clockwise (CCW) indicating actuation direction.

### Supplementary Figures

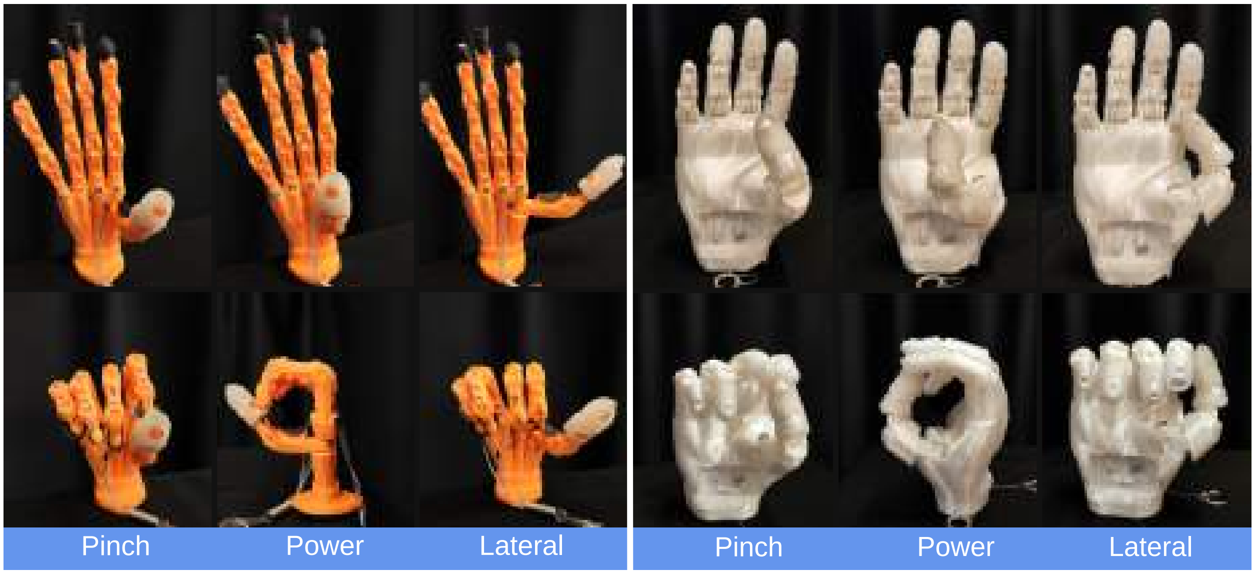


Figure S1: Representative grasp types with the Bionic Skeletal Hand (BSH). From left to right: pinch, power, and lateral grasps. The BSH achieves these grasp strategies by curling elongated fingers into a minimized palm with a passively oriented thumb.

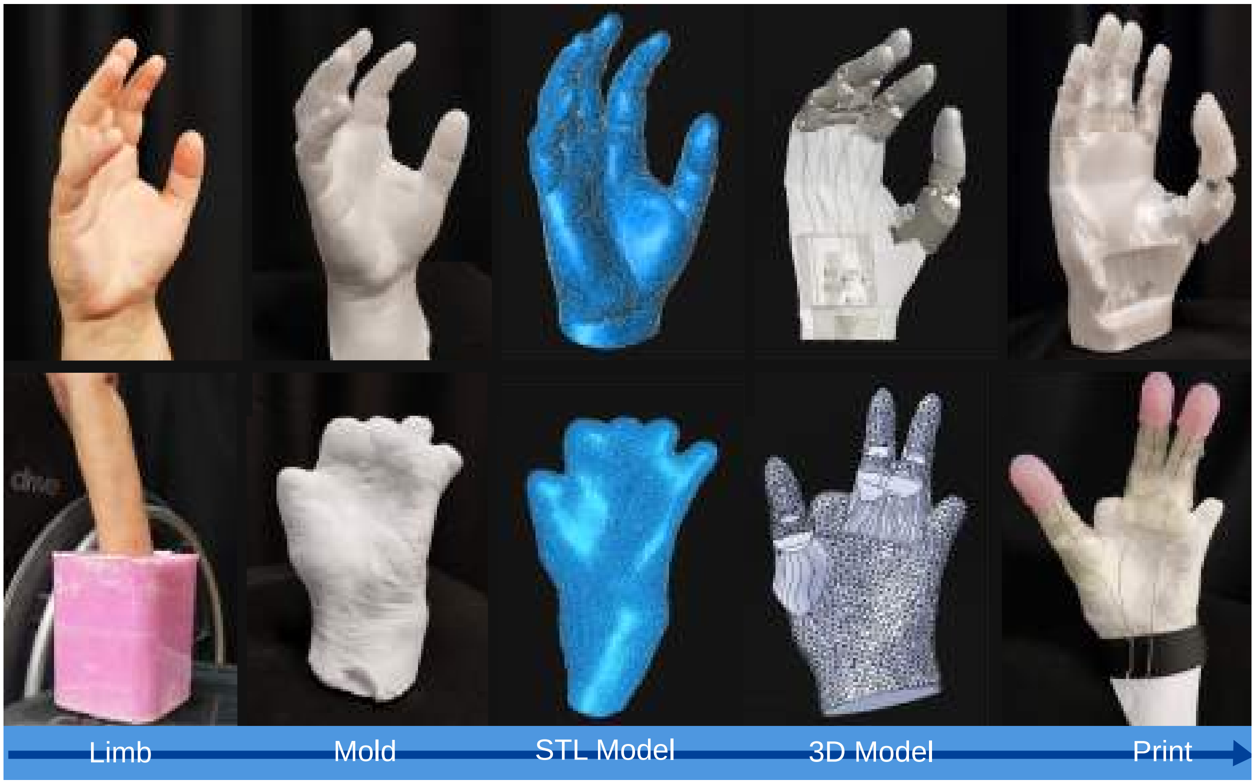


Figure S2: Five-step overview of the Human-Inspired Actuator Modeling and Reconstruction (HAMR) process applied to both a full-hand prosthetic end effector and a partial-hand fitting. The top row illustrates the HAMR workflow used to create the Tendon-Actuated Prosthetic Hand (TAPH), while the bottom row illustrates the same process applied to a flexible, 3D-printed fitting for a partial-hand prosthesis. From left to right, the steps include: (1) limb molding, (2) plaster casting, (3) 3D scanning, (4) CAD-based model reconstruction, and (5) additive manufacturing.

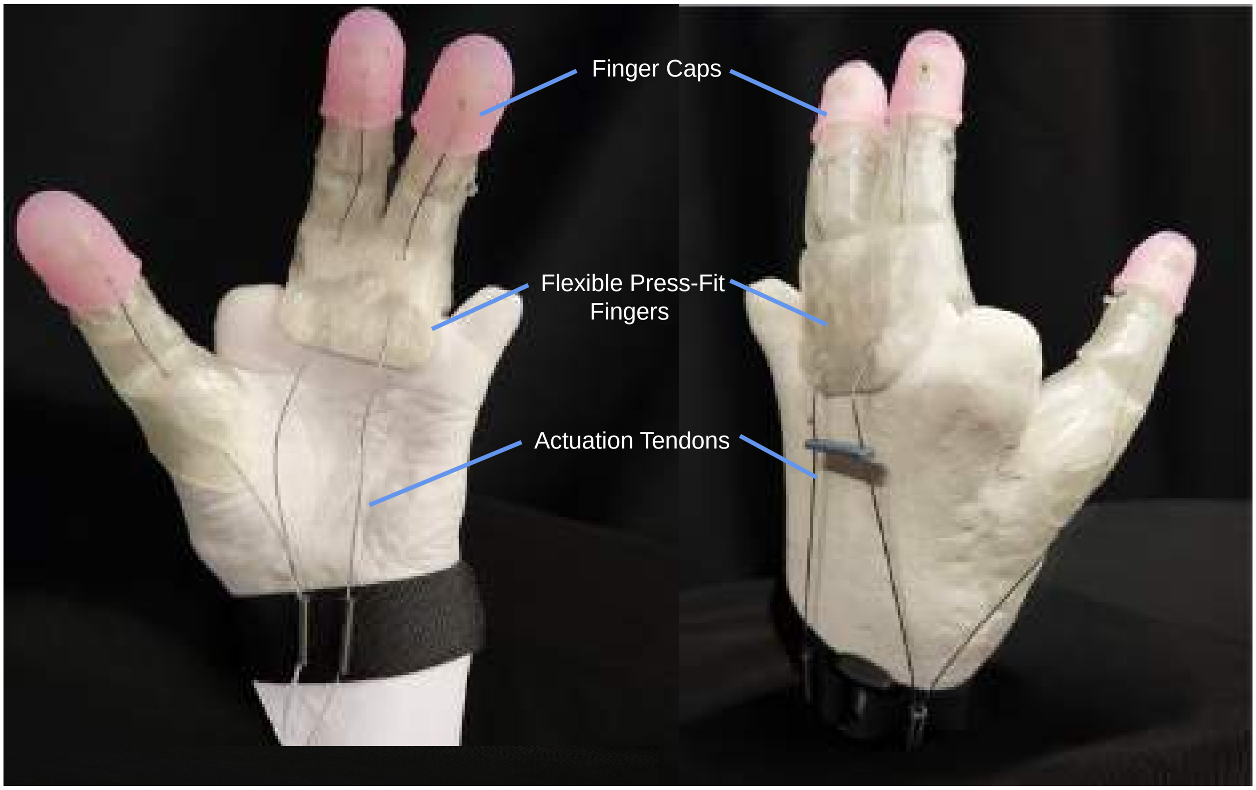


Figure S3: The multi-material custom partial hand fittings feature flexible press-fit bases, and rigid tendon-actuated finger joints.
